# Predictors to use mobile apps for monitoring COVID-19 symptoms and contact tracing: A survey among Dutch citizens

**DOI:** 10.1101/2020.06.02.20113423

**Authors:** Stephanie Jansen– Kosterink, Marian Hurmuz, Marjolein den Ouden, Lex van Velsen

**Affiliations:** Roessingh Research and Development, eHealth group, Enschede, the Netherlands; University of Twente, Faculty of Electrical Engineering, Mathematics and Computer Science, Telemedicine group, Enschede, the Netherlands; Saxion University of Applied Sciences, Research Center of Technology, Health & Care, Enschede, The Netherlands; ROC van Twente, Research Center Care and Technology, Hengelo, The Netherlands

**Keywords:** COVID-19, eHealth, mHealth, contact tracing, symptom management, intention to use

## Abstract

**Introduction:** eHealth applications have been recognized as a valuable tool to reduce COVID-19’s effective reproduction number. In this paper, we report on an online survey among Dutch citizens with the goal to identify antecedents of acceptance of a mobile application for COVID-19 symptom recognition and monitoring, and a mobile application for contact tracing.

**Methods:** Next to the demographics, the online survey contained questions focussing on perceived health, fear of COVID-19 and intention to use. We used snowball sampling via posts on social media and personal connections. To identify antecedents of acceptance of the two mobile applications we conducted multiple linear regression analyses.

**Results:** In total, 238 Dutch adults completed the survey. Almost 60% of the responders were female and the average age was 45.6 years (SD±17.4). For the symptom app, the final model included the predictors age, attitude towards technology and fear of COVID-19. The model had an R^2^ of 0.141. The final model for the tracing app included the same predictors and had an R^2^ of 0.156. The main reason to use both mobile applications was to control the spread of the COVID-19 virus. Concerns about privacy was mentioned as the main reason not to use the mobile applications.

**Discussion:** Age, attitude towards technology and fear of COVID-19 are important predictors of the acceptance of COVID-19 mobile applications for symptom recognition and monitoring and for contact tracing. These predictors should be taken into account during the development and implementation of these mobile applications to secure acceptance.

## 1 Introduction

It is spring 2020 and the COVID-19 pandemic has the world in its grip. Infection with COVID-19 can lead to a simple cold or no symptoms at all, while it can also rapidly develop into a life threatening disease, especially for patients with existing cardiovascular problems, obesity, or diabetes [1]. Death by COVID-19 is most often the result of massive alveolar damage and progressive respiratory failure [2]. Treatment of a severe COVID-19 infection can involve admission to the Intensive Care (IC) unit of a hospital, where the patient may be kept in a state of sleep and may be supported via artificial respiration. A stay at the IC may take weeks, resulting in high pressure on the availability of IC beds. In order to hamper the spread of COVID-19 and to manage the IC capacity, many countries have applied a lockdown strategy for their citizens [3]. At the moment of writing this article (May 2020), the spread of the virus is declining in Europe, and countries are loosening the restrictions, like lockdown they imposed upon their citizens. In order to control the spread of COVID-19 after a lockdown, and to minimize the effective reproduction number of the disease, several measures can be applied, of which social distancing, combined with aggressive case-finding and isolation seem to be the most effective [4].

eHealth applications have been recognized as a valuable tool for supporting symptom recognition and monitoring [5], for contact tracing [6], and ultimately, for reducing COVID-19’s effective reproduction number by means of timely intervention. In short, a contact tracing app would record a citizen’s contacts with other people via Bluetooth technology and, in the case of a COVID-19 infection, will warn the persons that the index patient recently had contact with so that they can apply self-isolation and be attentive for any COVID-19 symptoms. However, for such applications to be effective, high uptake among the population is necessary. For the case of a tracing application, it has been estimated that 56% of a country’s population should use the application to suppress the epidemic [7]. It is therefore crucial that the design of these applications and the implementation strategies that accompany them take the factors that affect acceptance into account.

The factors that determine acceptance of COVID-19 apps are largely unknown [8]. The exception here is privacy. Since the first plans of governments to implement these technologies, a fierce public debate has erupted on whether or not large scaling tracing of contacts for this goal is an unacceptable breach of privacy or not. While the issue of privacy has been recognized as an important antecedent of acceptance of mobile health applications [9], the unique and disturbing situation that the COVID-19 pandemic places us in, makes it difficult to apply existing models and frameworks for eHealth acceptance. The Dutch government wants to develop and implement two mobile applications to prevent the spread of the COVID-19 virus and support Dutch municipal health services. In this paper, we report on an online survey among Dutch citizens with the goal to identify antecedents of acceptance of 1) a mobile application for COVID-19 symptom recognition and monitoring, and 2) a mobile application for contact tracing.

## 2 Methods

To identify antecedents of acceptance of a mobile application for COVID-19 symptoms recognition and monitoring (hereafter: symptom app), and a mobile application for contact tracing (hereafter: tracing app), an online survey was developed, tested and distributed among Dutch citizens. This study did not require formal ethical approval (as ruled by CMO Arnhem Nijmegen, file number: 2020-6628). At the beginning of the survey, participants were asked for consent to use their data for research purposes.

### 2.1 Survey

The online survey (see appendix 1) consisted of four parts. The first part included questions on demographics, the second part contained questions related to perceived health, the third part consisted of questions related to the fear of a COVID-19 infection, and the final part included questions to assess the intention to use the two suggested mobile applications. In April 2020, the Dutch government announced plans to develop and implement two mobile applications for preventing the spread of the COVID-19 virus. However, the exact design of these applications remained unknown at this time. Therefore, we introduced both mobile applications in the survey via a short description of their general aim. We pre-tested the survey with 14 Dutch citizens to improve legibility.

#### 2.1.1 Demographics

We assessed gender, age, smartphone use, educational level (student, primary school, secondary school, high school, bachelor’s degree / University / PhD), work status (unemployed and searching for work, not able to work due to illness, volunteer work, part-time work, full-time work, retired, student), income level (below average wages, average wages, above average wages) and living status (living alone, living together, other). We assessed the participants’ attitude towards technology, using the Personal Innovativeness in the Domain of Information Technology scale by Agarwal & Prasad, 1998 [10], consisting of four statements and accompanied by a five-point Likert scale (ranging from 1 (strongly disagree) to 5 (strongly agree)). Finally, we also asked whether participants were (once) infected with COVID-19. The answer options for this question were: Yes, In doubt, or No.

#### 2.1.2 Perceived health

To assess perceived health, we asked participants to complete three questions. These questions were used previously to assess perceived health among Dutch citizens [11]. These questions/statements were: 1. How would you describe your health?; 2. How concerned are you about your health?; and 3. I am ill more often than other people of the same age and sex. These were accompanied by a five-point Likert scale ranging from 1 (bad, not concerned and totally disagree, respectively) to 5 (excellent, very concerned and totally agree, respectively).

#### 2.1.3 Fear of COVID-19

The participants’ fear of a COVID-19 infection was assessed by means of four questions related to this topic.

- Have you been concerned about the outbreak of the COVID-19 virus in recent weeks? [five-point Likert scale, ranging from 1 (not at all concerned) to 5 (extremely concerned)];
- How often did you think of the outbreak of the COVID-19 virus in recent weeks? [five-point Likert scale, ranging from 1 (never) to 5 (always)];
- How afraid were you of the outbreak of the COVID-19 virus in recent weeks? [five-point Likert scale, ranging from 1 (not afraid at all) to 5 (very afraid)];
- How afraid are you of getting sick from the COVID-19 virus? [five-point Likert scale, ranging from 1 (not afraid at all) to 5 (very afraid)].

#### 2.1.4 Intention to use

Finally, participants were asked to rate their intention to use the two mobile applications: 1) a symptom app, and 2) a tracing app. The statements for the construct intention to use were based on Van Velsen et al., 2015 [12]. All three questions were accompanied by a five-point Likert scale ranging from 1 (strongly disagree) to 5 (strongly agree). Next to these closed questions, respondents were also asked what the main reasons were to ‘use’ and ‘not to use’ the mobile applications.

### 2.2 Survey distribution

Distribution of the survey started on April 15, 2020. Participants were eligible if they were 18 years of age or older. We used a snowball sampling via posts on social media (LinkedIn, Twitter and Facebook) and personal connections. Next to this, we recruited participants via a Dutch panel of older adults that indicated they were interested in participating in research on the topic of eHealth. The survey was closed on April 30, 2020. Due to the method of recruitment, a response rate could not be calculated.

### 2.3 Analyses

Data were analysed by using SPSS, version 19. Descriptive statistics were performed for all outcomes. Cronbach’s alphas were calculated to assess internal consistency for attitude towards technology, perceived health, fear of COVID-19 and intention to use. Next, survey scores were interpreted for these factors as being negative (score 1 or 2), neutral (score of 3), or positive (score 4 or 5). Via a paired t-test, the difference in intention to use score between both mobile applications was tested. To identify antecedents of acceptance of 1) a symptom app, and 2) a tracing app, we conducted multiple linear regression analyses (backward model analyses). The intention to use each app was used as the dependent variable. The independent variables were those demographic characteristics and factors that (borderline) significant correlated (Pearson Correlation cut-off level p≤0.10) with the dependent variable. For the paired t-test and regression analyses, the level of significance was set at p < 0.05.

## 3 Results

In total, 238 Dutch citizens completed the survey. Fifteen responders only completed the intention to use survey of a tracing app as this app was presented first and these responders stopped with the survey after these questions. Almost 60% of the responders were female and the average age was 45.6 years (SD±17.4). Only five responders (2.1%) did not own a smartphone and almost 75% claimed that they carried their smartphone with them for most of the day. The internal consistence of the attitude towards technology scale was good (Cronbach’s Alpha = 0.85). Most responders (73.9%) had a moderate attitude towards technology. Only three responders (1.3%) claimed to be infected with COVID-19. All demographic characteristics are presented in table 1.

**Table 1:** Responders’ demographics.

| Demographic (n=238) |  |  |
| --- | --- | --- |
| Gender | Male | 40.8% |
|  | Female | 59.2% |
| Age | | 45.6 ($SD \pm 17.4$) years |
| Smartphone | Yes | 97.9% |
|  | No | 2.1% |
| Carry smartphone with you? | Always | 74.8% |
|  | Sometimes | 23.1% |
|  | Never | 2.1% |
| Education level | Student | 6.7% |
|  | Primary school | 0.8% |
|  | Secondary school | 5.9% |
|  | High school | 23.9% |
|  | Bachelor's degree / University / PhD | 62.6% |
| Work status | Unemployed and searching for work | 1.3% |
|  | Not able to work due to illness | 3.4% |
|  | Volunteer work | 0.8% |
|  | Part-time work | 31.5% |
|  | Full-time work | 34.4% |
|  | Retired | 18.1% |
|  | Student | 10.5% |
| Income level | Below average wages | 31.9% |
|  | Average wages | 39.1% |
|  | Above average wages | 29% |
| Living status | Living alone | 14.3% |
|  | Living together | 80.3% |
|  | Other | 5.5% |
| Attitude towards technology |  | 3.2 (SD±0.78) [1-5] |
|  | Low (1-2) | 1.3% |
|  | Moderate (3) | 73.9% |
|  | High (4-5) | 24.8% |
| COVID-19 infection | Yes | 1.3% |
|  | In doubt | 18.5% |
|  | No | 80.3% |

### 3.1 Fear of a COVID-19 infection

The internal consistency of the four items in this scale was acceptable to good (Cronbach’s Alpha = 0.78). The mean score on this topic was 3.3 (SD±0.68). The majority of the responder’s opinion on this topic was neutral (80.7%) and 16% of the responders were afraid for a COVID-19 infection. Only a few responders (3.4%) were not afraid (table 2).

### 3.2 Perceived health

For the three items to assess the perceived health of the responders the internal consistence was acceptable (Cronbach’s Alpha = 0.69). The mean score on this scale was 3.8 (SD±0.68). Most respondents were positive about their health (58.4%).

### 3.3 Intention to use

The intention to use was assessed for the symptom app and the tracing app. For both scales, internal consistency was excellent; Cronbach’s Alpha symptom app = 0.96 and Cronbach’s Alpha tracing app = 0.96. For both apps, the majority’s intention to use was neutral (see table 2). However, an additional paired t-test indicated that there is a significant difference in the scores on intention to use for the symptom app (M=3.38, SD±1.07, n=223) and the tracing app (M=3.27, SD±1.13, n=223); t(222)=-2.598 and p=0.01. Indicating that the responders were more willing to use a mobile application for COVID-19 symptom recognition and monitoring compared to a mobile application for contact tracing.

**Table 2:** Descriptive statistics and internal consistency of scales.

| Scale | Number of items | Cronbach's Alpha | Mean (SD) | Positive | Neutral | Negative |
| --- | --- | --- | --- | --- | --- | --- |
| Fear of COVID-19 | 4 | 0.78 | 3.3 (SD±0.68) | 16% | 80.7% | 3.4% |
| Perceived health | 3 | 0.69 | 3.8 (SD±0.68) | 58.4% | 40.8% | 0.8% |
| Symptom app (n=223) |  |  |  |  |  |  |
| Intention to use | 3 | 0.96 | 3.38<br>(SD±1.07) | 45.3% | 45.3% | 9.4% |
| Tracing app (n=238) |  |  |  |  |  |  |
| Intention to use | 3 | 0.96 | 3.27<br>(SD±1.14) | 41.2% | 45.4% | 13.4% |

### 3.4 Correlations

The intention to use a symptom app is related to income level (r=0.132, p<0.05), attitude towards technology (r=0.220, p<0.01) and fear of COVID-19 (r=-0.291, p<0.01). The intention to use a tracing app is related to age (r=0.135, p=0.04), attitude towards technology (r=0.223, p<0.01) and fear of COVID-19 (r=-0.303, p<0.01). Based on these outcomes the independent variables within the linear regression analysis were: age, income level, attitude towards technology, fear of COVID-19 and perceived health. Table 3 provides an overview of the correlations between all demographics and factors, and the intention to use.

**Table 3:** Outcome Pearson Correlation.

|  | Intention to use<br>symptom app (n=223) | Intention to use<br>tracing app (n=238) |
| --- | --- | --- |
| Gender | $r=-0.056$<br>$p=0.41$ | $r=-0.147$<br>$p=0.23$ |
| Age | $r=0.126$<br>$p=0.06$ | $r=0.135^*$<br>$p=0.04$ |
| Education level | $r=0.21$<br>$p=0.76$ | $r=0.018$<br>$p=0.79$ |
| Work status | $r=0.072$<br>$p=0.28$ | $r=0.033$<br>$p=0.62$ |
| Income level | $r=-0.132^*$<br>$p=0.05$ | $r=0.124$<br>$p=0.06$ |
| Living status | $r=0.083$<br>$p=0.218$ | $r=0.060$<br>$p=0.353$ |
| Attitude towards<br>technology | $r=0.220^*$<br>$p<0.01$ | $r=0.223^*$<br>$p<0.01$ |
| Fear of COVID-19 | $r=-0.291^*$<br>$p<0.01$ | $r=-0.303^*$<br>$p<0.01$ |
| Perceived health | $r=-0.088$<br>$p=0.19$ | $r=-0.119$<br>$p=0.07$ |

\*Correlation is significant at the 0.05 level (2-tailed).

#### 3.4.1 Linear regression

A multiple linear regression analysis was conducted to predict the intention to use a symptom app based on age, income level, attitude towards technology, fear of COVID-19 and perceived health. The final model included the predictors attitude towards technology, fear of COVID-19, and age. The model has an R^2^ of 0.141. It contains three factors that affect the intention to use, but only two of them are significant predictors:

- Fear of COVID-19, β=0.272, t=4.305, p<0.001;
- Attitude towards technology, β=0.222, t=3.532, p<0.01;
- Age, β=0.107, t=1.691, not significant.

Another multiple linear regression analysis was conducted to predict the intention to use a tracing app based on age, income level, attitude towards technology, fear of COVID-19 and perceived health. The final model included the predictors attitude towards technology, fear of COVID-19, and age. The model has a R^2^ of 0.155. Intention to use is predicted by:

- Fear of COVID-19, β=0.286, t=4.742, p<0.001;
- Attitude towards technology, β=0.230, t=3.815, p<0.001;
- Age, β=0.128, t=2.104, p<0.05.

### 3.5 Main reason to use the mobile applications

An overview of all reasons the responders brought forth for using both mobile applications is presented in table 4. The main reason (28.4%) for responders to use the symptom app, was to control the spread of the COVID-19 virus. In addition, respondents were willing to use this mobile application to monitor own complaints (19.0%) and to gain more insight into the spread and symptoms of the COVID-19 virus (16.4%).

The main reason to use a tracing app was also to control the spread of the COVID-19 virus (30.6%). Next to this, respondents were willing to use this mobile application to gain more insight into the spread and symptoms of the COVID-19 virus (23.1%) and for one’s own health (12.9%).

**Table 4:**
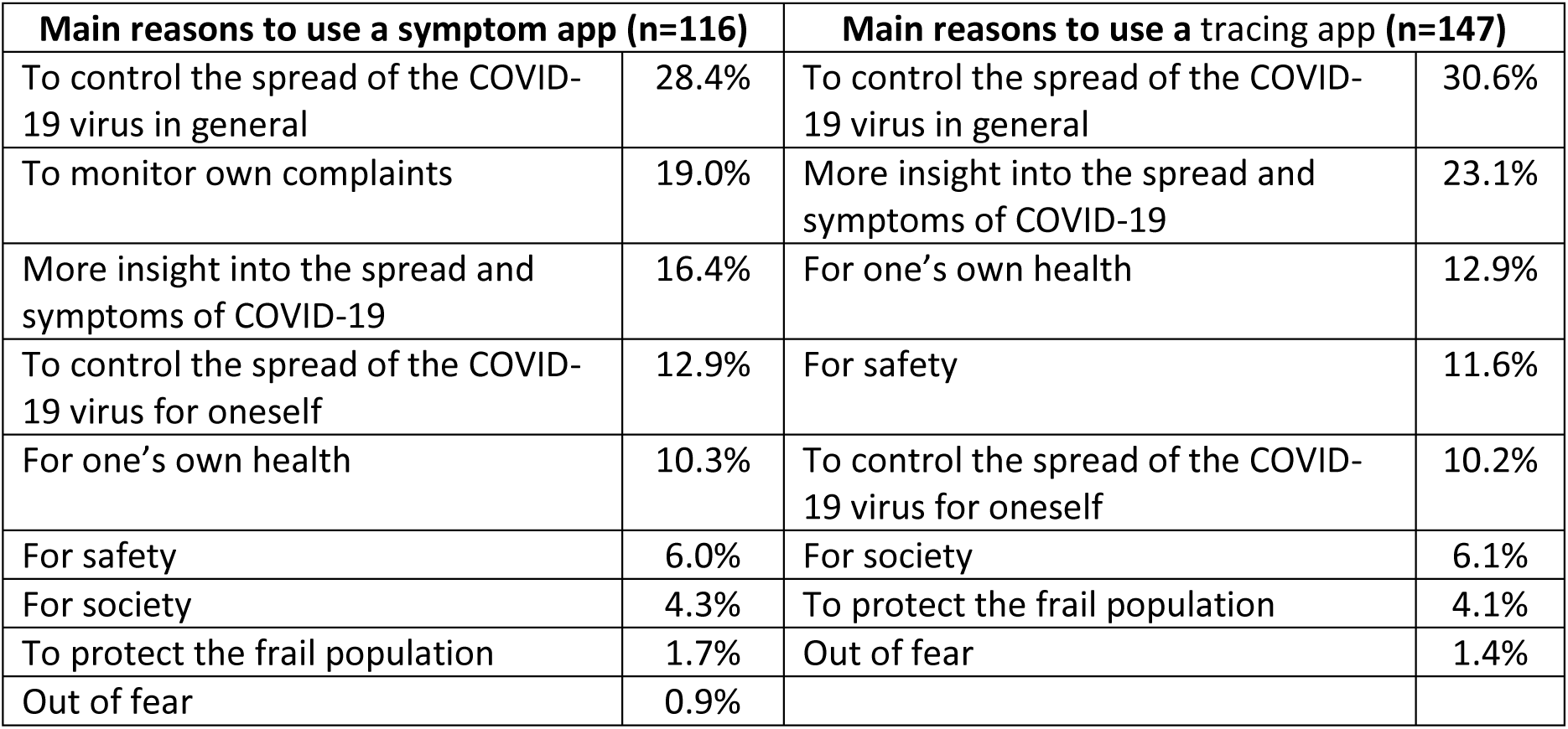
Overview of the main reasons to use the two mobile applications.

| Main reasons to use a symptom app (n=116) |  | Main reasons to use a tracing app (n=147) |  |
| --- | --- | --- | --- |
| To control the spread of the COVID-19 virus in general | 28.4% | To control the spread of the COVID-19 virus in general | 30.6% |
| To monitor own complaints | 19.0% | More insight into the spread and symptoms of COVID-19 | 23.1% |
| More insight into the spread and symptoms of COVID-19 | 16.4% | For one's own health | 12.9% |
| To control the spread of the COVID-19 virus for oneself | 12.9% | For safety | 11.6% |
| For one's own health | 10.3% | To control the spread of the COVID-19 virus for oneself | 10.2% |
| For safety | 6.0% | For society | 6.1% |
| For society | 4.3% | To protect the frail population | 4.1% |
| To protect the frail population | 1.7% | Out of fear | 1.4% |
| Out of fear | 0.9% |  |  |

### 3.6 Main reason <u>not</u> to use the mobile applications

An overview of the reasons not to use the mobile applications is presented in table 5. For both mobile applications, privacy was mentioned as the main reason (symptom app=55.7% and tracing app=64.8%) not to use the mobile applications. Other reasons for not using the mobile applications were the expected usefulness of the application (symptom app=23.5% and tracing app=13.4%) and a fear of becoming over aware of the situation and its potential consequences, leading to unnecessary stress (symptom app=7.8% and tracing app=11.3%).

**Table 5:** Overview of the main reasons not to use the two mobile applications.

| Main reasons to use a symptom app (n=113) |  | Main reasons not to use a tracing app (n=142) |  |
| --- | --- | --- | --- |
| Privacy / not willing to share information with government | 55.7% | Privacy / not willing to share information with government | 64.8% |
| Doubting usefulness | 23.5% | Doubting usefulness | 13.4% |
| Over awareness / stress | 7.8% | Over awareness / stress | 11.3% |
| Doubting ease of use | 4.3% | No (compatible) phone | 4.2% |
| Doubting security | 4.3% | Doubting security | 2.1% |
| No (compatible) phone | 1.7% | Doubting ease of use | 2.1% |
| The fear the use of the app will be forced by government | 0.9% | The fear the use of the app will be forced by government | 2.1% |

## 4 Discussion

The aim of this paper was to identify antecedents of acceptance of 1) a mobile application for COVID-19 symptom recognition and monitoring, and 2) a mobile application for contact tracing among Dutch citizens by means of an online survey. A large group of the Dutch citizens (45.3%) is willing to use a mobile application for COVID-19 symptom recognition and monitoring. The main reasons to use this mobile application are: 1. To control the spread of the COVID-19; 2. To monitor their own complaints; and 3. To gain more insight into the spread and symptoms of the COVID-19 virus. For the case of a mobile application for COVID-19 contact tracing, 41.2% of the Dutch adults appears to be willing to use this mobile application. The main reasons for use are: 1. To control the spread of the COVID-19 virus; 2. To gain more insight into the spread and symptoms of the COVID-19 virus; and 3. For their own health. Privacy, doubting the usefulness of the mobile application and a fear of becoming over aware of the situation and its potential consequences, leading to unnecessary stress are the main reasons not to use the mobile applications. For both mobile applications age, attitude towards technology and fear of COVID-19 are antecedents of acceptance.

It is difficult to relate our findings to the existing literature, as technology acceptance studies have not focused on mobile applications to be used during a pandemic, and insights on factors that determine the acceptance of COVID-19 related mobile applications are lacking [8]. In general, age and attitude towards technology are widely-acknowledged antecedents of acceptance. For age there is evidence that older age is associated with lower level of acceptance of mobile applications [13]. Previous results also indicated that attitude towards technology is an important antecedent of acceptance of mobile applications [13, 14]. The degree to which an individual is willing to try out any new mobile application is related to the intention to use [13].

Our results show that fear of COVID-19 is the most important COVID-19-related factor that predicts acceptance of mobile applications to deal with the COVID-19 pandemic. Since it is difficult to translate this fear into technology design, this finding needs to be seen in a bigger picture. Public health campaigns during the COVID-19 epidemic will need to educate citizens about the dangers of COVID-19 (personally and for society as a whole), and should then offer downloading COVID-19 mobile applications as a personal strategy to deal with this fear. Next, the positive attitude towards technology that precedes a decision to download a COVID-19 app should be taken into consideration when using these innovations. The end-user population might be skewed towards those with interest in technology (traditionally these are younger, highly-educated men [15]) which can create a use divide, and thus, a health divide in society. Measures should be installed to support those groups in society that are not, by nature, technically interested, like promotional stalls in the community and diverse channels of user support.

The following four limitations should be taken into account for this study. First, the selection bias of our sample. Given the results the education level of our sample is higher compared to the general population. In addition, most respondents had a moderate attitude towards technology. This could be explained by our way to distribute the survey by snowball sampling. Second, for our analysis the power of our sample was sufficient. However, a larger sample would improve the generalizability of our outcomes as mainly Dutch citizens from the eastern part of the Netherlands (87% of our sample) completed our survey. Third, in our survey the two mobile applications are introduced by means of a short description of their general aim. It is unclear if this description was sufficient for the responders to understand to purpose of both mobile applications. Fourth, the explained variance of both our models is relatively low. Normally, in studies such as these, this number is boosted by including the predictors ease of use and perceived usefulness. However, since including these factors leads to little practical results (concluding that the applications should be easy to use is a given and does not inform design), the identification COVID-19-related factors is an important extension of existing technology acceptance models.

## 5 Conclusion

This study is the first to determine the factors related to the acceptance of COVID-19 apps. Age, attitude towards technology and fear of COVID-19 are important predictors of the acceptance of COVID-19 mobile applications for symptom recognition and monitoring and for contact tracing. These predictors should be taken into account during the development and implementation of these mobile applications to secure acceptance.

## 6 Authors’ contributions

The survey was developed by SJK, MH and LvV. Statistical analyses were performed by SJK and LvV. All authors were involved in the distribution of the survey and participated in drafting the article and revising it critically for important intellectual content.

## Data Availability

The data that support the findings of this study are available on request from the corresponding author

## 7 Acknowledgments

Not applicable

## 8 Summary table

| What was already known on the topic: | What this study added to our knowledge: |
| --- | --- |
| eHealth applications have been recognized as a valuable tool to reduce COVID-19's effective reproduction number by means of timely intervention<br>The factors that determine acceptance of COVID-19 apps are unknown<br>eHealth could be a solution to rapidly cope with health demands during a pandemic as an addition to the health care infrastructure | 41-45% of the Dutch adults are willing to use mHealth to prevent the spread of COVID-19<br>Age, attitude towards technology and fear of COVID-19 predict the intention to use mHealth to prevent the spread of COVID-19<br>The main reasons to use this application are to control the spread of the COVID-19 virus for the general purpose, to monitor their own complaints and to gain more insight into the spread and symptoms of the COVID-19 virus<br>Privacy, doubting the usefulness of the mHealth application and fear of over awareness leading to unnecessary stress |
|  | are the main reasons not to use mHealth to prevent the spread of COVID-19 |

## 10 Appendix 1

**Table 6:** Survey questions and answer options in Dutch and English. (D= demographic questions / C= fear of COVID-19 questions / H= perceived health questions / TAM-BI= behavioural intention)

|  |  | Dutch |  | English |  |
| --- | --- | --- | --- | --- | --- |
| D | 1 | Wat is uw geslacht? | o Man<br>o Vrouw | What is your gender? | o Men<br>o Women |
| D | 2 | Wat is uw leeftijd? |  | What is your age? |  |
| D | 3 | Wat zijn de 4 cijfers van uw postcode? |  | What are the 4 digits of your zip code? |  |
| D | 4 | Heeft u een smartphone? | o Ja<br>o Nee | Do you have a smartphone? | o Yes<br>o No |
| D | 5 | Draagt u uw smartphone de hele dag bij u? | o Nooit<br>o Soms<br>o Altijd | Do you carry your smartphone with you all day? | o Never<br>o Sometimes<br>o Always |
| D | 6 | Wat is de hoogste opleiding die u heeft afgerond? | o Basisschool<br>o Lbo, mavo, vmbo<br>o Mbo, havo, vwo<br>o Hbo, wo<br>o Ik studeer nog | What is the highest level of education you have completed? | o Primary school<br>o vocation education<br>o vocational education<br>o higher education<br>o I am still studying |
| D | 7 | Welke van de volgende categorieën beschrijft uw werkstatus het best? | o Werkloos, op zoek naar werk<br>o Werkloos, niet op zoek naar werk<br>o Part-time werkzaam<br>o Full-time werkzaam<br>o Gepensioneerd<br>o Door ziekte niet de mogelijkheid om te werken<br>o Student<br>o Vrijwilligerswerk | Which of the following categories best describes your work status? | o Unemployed, looking for work<br>o Unemployed, not looking for work<br>o Working part-time<br>o Working full-time<br>o Retired<br>o Not able to work due to illness<br>o Student<br>o Volunteering |
| D | 8 | Wat is uw gemiddelde inkomen? (Modaal inkomen = €36.000 bruto per jaar) | o Beneden modaal<br>o Rond modaal<br>o Boven modaal | What is your average income? (Average income = € 36,000 gross per year) | o Below average<br>o Around average<br>o Above average |
| D | 9 | Bent u alleen of woont u samen? | o Alleenstaand<br>o Samenwonend<br>o Anders | Are you single or do you live together? | o Single<br>o Living together<br>o Otherwise |
| D | 10 | Hoe denkt u over nieuwe technologieën in het algemeen? Vink voor elke stelling het antwoord aan dat het beste bij u past. |  | How do you feel about new technologies in general? For each statement, tick the answer that suits you best. |  |
|  |  | a. Als ik hoor over een nieuwe technologie, kijk ik ernaar uit om dat uit te proberen | o Zeer mee oneens<br>o Mee oneens<br>o Neutraal<br>o Mee eens<br>o Zeer mee eens | a. When I hear about a new technology, I look forward to trying it out | o Strongly disagree<br>o I disagree<br>o Neutral<br>o Agree<br>o Strongly agree |
|  |  | b. Vergeleken met de mensen in mijn omgeving ben ik meestal een van de eersten die nieuwe technologieën uitprobeert | o Zeer mee oneens<br>o Mee oneens<br>o Neutraal<br>o Mee eens<br>o Zeer mee eens | b. Compared to the people around me, I'm usually one of the first to try out new technologies | o Strongly disagree<br>o I disagree<br>o Neutral<br>o Agree<br>o Strongly agree |
|  |  | c. In het algemeen aarzel ik om nieuwe technologieën uit te proberen | ☐ Zeer mee oneens<br>☐ Mee oneens<br>☐ Neutraal<br>☐ Mee eens<br>☐ Zeer mee eens | c. In general, I hesitate to try out new technologies | ☐ Strongly disagree<br>☐ I disagree<br>☐ Neutral<br>☐ Agree<br>☐ Strongly agree |
|  |  | d. Ik probeer graag nieuwe technologieën uit | ☐ Zeer mee oneens<br>☐ Mee oneens<br>☐ Neutraal<br>☐ Mee eens<br>☐ Zeer mee eens | I like to try out new technologies | ☐ Strongly disagree<br>☐ I disagree<br>☐ Neutral<br>☐ Agree<br>☐ Strongly agree |
| D | 11 | Bent u al besmet geraakt met het COVID-19 virus? | ☐ Ja<br>☐ Nee<br>☐ Ik twijfel | Have you already been infected with the COVID-19 virus? | ☐ Yes<br>☐ No<br>☐ I doubt |
| C | 12 | Was u de afgelopen weken bezorgd over de uitbraak van het COVID-19 virus? | ☐ Helemaal niet bezorgd<br>☐ Een klein beetje bezorgd<br>☐ Nogal bezorgd<br>☐ Bezorgd<br>☐ Heel erg bezorgd | Have you been concerned about the outbreak of the COVID-19 virus in recent weeks? | ☐ Not at all concerned<br>☐ Slightly concerned<br>☐ Somewhat concerned<br>☐ Moderately concerned<br>☐ Extremely concerned |
| C | 13 | Hoe vaak dacht u de afgelopen weken aan de uitbraak van het COVID-19 virus? | ☐ Nooit<br>☐ Zelden<br>☐ Soms<br>☐ Vaak<br>☐ Altijd | How often did you think of the outbreak of the COVID-19 virus in recent weeks? | ☐ Never<br>☐ Rarely<br>☐ Sometimes<br>☐ Often<br>☐ Always |
| C | 14 | Hoe bang was u de afgelopen weken voor de uitbraak van het COVID-19 virus? | ☐ Helemaal niet bang<br>☐ Een klein beetje bang<br>☐ Nogal bang<br>☐ Bang<br>☐ Heel erg bang | How scared were you of the outbreak of the COVID-19 virus in recent weeks? | ☐ Not afraid at all<br>☐ A little bit afraid<br>☐ Quite afraid<br>☐ Scared<br>☐ Very scared |
| C | 15 | Hoe bang bent u om ziek te worden van het COVID-19 virus? | ☐ Helemaal niet bang<br>☐ Een klein beetje bang<br>☐ Nogal bang<br>☐ Bang<br>☐ Heel erg bang | How afraid are you of getting sick from the COVID-19 virus? | ☐ Not afraid at all<br>☐ A little bit afraid<br>☐ Quite afraid<br>☐ Scared<br>☐ Very scared |
| H | 16 | Hoe zou u uw gezondheid omschrijven? | ☐ Slecht<br>☐ Matig<br>☐ Goed<br>☐ Zeer goed<br>☐ Uitstekend | How would you describe your health? | ☐ Bad<br>☐ Poor<br>☐ Good<br>☐ Very good<br>☐ Excellent |
| H | 17 | Hoe bezorgd bent u over uw gezondheid? | ☐ Helemaal niet bezorgd<br>☐ Een klein beetje bezorgd<br>☐ Nogal bezorgd<br>☐ Bezorgd<br>☐ Heel erg bezorgd | How concerned are you about your health? | ☐ Not at all concerned<br>☐ Slightly concerned<br>☐ Somewhat concerned<br>☐ Moderately concerned<br>☐ Extremely concerned |
| H | 18 | Ik ben vaker ziek dan andere mensen van dezelfde leeftijd en hetzelfde geslacht. | ☐ Zeer mee oneens<br>☐ Mee oneens<br>☐ Neutraal<br>☐ Mee eens<br>☐ Zeer mee eens | I am sick more often than other people of the same age and gender. | ☐ Strongly disagree<br>☐ I disagree<br>☐ Neutral<br>☐ Agree<br>☐ Strongly agree |
| TAM | 19 | Hieronder krijgt u stellingen die gaan over uw verwachting over de app waarmee in kaart wordt gebracht met wie een besmet persoon in contact is geweest. Vink voor elke stelling het antwoord aan wat het best aan uw verwachting voldoet. |  | Below you will find statements about your expectation about the app to determine if an infected person has been contacted with others. For each statement, tick the answer that best meets your expectations. |  |
| TAM-BI |  | a. Ik ben van plan deze App te gebruiken zo vaak als nodig is. | ☐ Zeer mee oneens<br>☐ Mee oneens<br>☐ Neutraal<br>☐ Mee eens<br>☐ Zeer mee eens | a. I plan to use this App as often as necessary. | ☐ Strongly disagree<br>☐ I disagree<br>☐ Neutral<br>☐ Agree<br>☐ Strongly agree |
|  |  | b. Als deze App beschikbaar zou zijn voor mij, zou ik deze absoluut gebruiken. | ☐ Zeer mee oneens<br>☐ Mee oneens<br>☐ Neutraal<br>☐ Mee eens<br>☐ Zeer mee eens | b. If this App were available to me, I would absolutely use it. | ☐ Strongly disagree<br>☐ I disagree<br>☐ Neutral<br>☐ Agree<br>☐ Strongly agree |
|  |  | c. Ik hoop dat deze App beschikbaar komt voor mij. | ☐ Zeer mee oneens<br>☐ Mee oneens<br>☐ Neutraal<br>☐ Mee eens<br>☐ Zeer mee eens | c. I hope this App becomes available to me. | ☐ Strongly disagree<br>☐ I disagree<br>☐ Neutral<br>☐ Agree<br>☐ Strongly agree |
|  | 20 | Wat is voor u de belangrijkste reden om gebruik te maken van deze App? |  | What is the main reason for you to use this App? |  |
|  | 21 | Wat is voor u de belangrijkste reden om geen gebruik te maken van deze App? |  | What is the main reason for you not to use this App? |  |
| TAM | 22 | Hieronder krijgt u stellingen die gaan over uw verwachting over de app om symptomen van u, als eventuele corona patiënt, te volgen. Vink voor elke stelling het antwoord aan wat het best aan uw verwachting voldoet. |  | Below you will find statements about your expectation about the app to track your symptoms as corona patient. For each statement, tick the answer that best meets your expectations. |  |
| TAM-BI |  | a. Ik ben van plan deze App te gebruiken zo vaak als nodig is. | ☐ Zeer mee oneens<br>☐ Mee oneens<br>☐ Neutraal<br>☐ Mee eens<br>☐ Zeer mee eens | a. I plan to use this App as often as necessary. | ☐ Strongly disagree<br>☐ I disagree<br>☐ Neutral<br>☐ Agree<br>☐ Strongly agree |
|  |  | b. Als deze App beschikbaar zou zijn voor mij, zou ik deze absoluut gebruiken. | ☐ Zeer mee oneens<br>☐ Mee oneens<br>☐ Neutraal<br>☐ Mee eens<br>☐ Zeer mee eens | b. If this App were available to me, I would absolutely use it. | ☐ Strongly disagree<br>☐ I disagree<br>☐ Neutral<br>☐ Agree<br>☐ Strongly agree |
|  |  | c. Ik hoop dat deze App beschikbaar komt voor mij. | ☐ Zeer mee oneens<br>☐ Mee oneens<br>☐ Neutraal<br>☐ Mee eens<br>☐ Zeer mee eens | c. I hope this App becomes available to me. | ☐ Strongly disagree<br>☐ I disagree<br>☐ Neutral<br>☐ Agree<br>☐ Strongly agree |
|  | 23 | Wat is voor u de belangrijkste reden om gebruik te maken van deze App? |  | What is the main reason for you to use this App? |  |
|  | 24 | Wat is voor u de belangrijkste reden om geen gebruik te maken van deze App? |  | What is the main reason for you not to use this App? |  |

